# Rapid and sustained decline in CXCL-10 (IP-10) annotates clinical outcomes following TNF-α antagonist therapy in hospitalized patients with severe and critical COVID-19 respiratory failure

**DOI:** 10.1101/2021.05.29.21258010

**Authors:** Hilal Hachem, Amandeep Godara, Courtney Schroeder, Daniel Fein, Hashim Mann, Christian Lawlor, Jill Marshall, Andreas Klein, Debra Poutsiaka, Janis L. Breeze, Raghav Joshi, Paul Mathew

## Abstract

**Background:** A feed-forward pathological signaling loop generated by TNFα and IFN-γ in inflamed lung tissue, driving CXCL-10 (IP-10) and CXCL-9 chemokine-mediated activated T-cell and monocyte/macrophage tissue recruitment, may define, sustain and amplify the inflammatory biology of lethal COVID-19 respiratory failure.

**Methods:** To assess TNFα-antagonist therapy, 18 hospitalized adults with hypoxic respiratory failure and COVID-19 pneumonia received single-dose infliximab-abda therapy 5mg/kg intravenously between April and December 2020. The primary endpoint was time to increase in oxygen saturation to fraction of inspired oxygen ratio (SpO2/FiO2) by ≥ 50 compared to baseline and sustained for 48 hours. Secondary endpoints included 28-day mortality, dynamic cytokine profiles (Human Cytokine 48-Plex Discovery Assay, Eve Technologies), secondary infections, duration of supplemental oxygen support and hospitalization.

**Findings:** Patients were predominantly in critical respiratory failure (15/18, 83%), male (14/18, 78%), above 60 years (median 63 yrs, range 31-80), race-ethnic minorities (13/18, 72%), lymphopenic (13/18, 72%), steroid-treated (17/18, 94%), with a median ferritin of 1953ng/ml. Sixteen patients (89%) met the primary endpoint within a median of 4 days, 15/18 (83%) recovered from respiratory failure, and 14/18 (78%) were discharged in a median of 8 days and were alive at 28-day follow-up. Deaths among three patients ≥ 65yrs age with pre-existing lung disease or multiple comorbidities were attributed to secondary lung infection. Mean plasma IP-10 levels declined sharply from 9183 pg/ml to 483 pg/ml at Day 3 and further to 146 pg/ml at Day 14/discharge. Significant declines in IFN-*γ*, TNFα, IL-27, CRP and ferritin were specifically observed at Day 3 whereas other cytokines were unmodified. IL-6 levels declined sharply among patients with baseline levels >10 pg/ml. Among 13 lymphopenic patients, six (46%) had resolution of lymphopenia by day 3, and 11 by day 14. CXCR3-ligand (IP-10 and CXCL-9) declines were strongly correlated among patients with lymphopenia reversal (Day 3, Pearson r: 0.98, p-value: 0.0006).

**Interpretation:** Consistent with a pathophysiological role of TNFα, the clinical and cytokine data indicate that infliximab-abda may rapidly abrogate pathological inflammatory signaling to facilitate clinical recovery in severe and critical COVID-19. Randomized studies are required to formally assess mortality outcomes. Funding: National Center for Advancing Translational Sciences

## Introduction

Preclinical and clinical evidence indicates that TNFα may fundamentally orchestrate the hyperinflammatory cytokine signature and adverse disease course of COVID-19.^1,2^ Elevated circulating levels of TNFα and its known regulatory targets including interleukin-6 (IL-6) and ferritin, have been correlated with indices of disease severity, including intensive care needs, multisystem organ failure and death.^3-5^ A marked quantitative reduction in lymphocytes and natural killer cells in severe COVID-19 illness is accompanied by markers of immune exhaustion, which are inversely associated with levels of IL-6, IL-10 and TNFα.^6^ An aberrant hyperimmune response triggered by synergy between TNFα and IFN-γ in inflamed lung tissue may underpin the generation of markedly elevated T-cell chemokines CXCL-10 (IP-10) and CXCL-9, associated with immune exhaustion and adverse clinical outcomes.^1,7,8^ TNFα has been previously implicated as a master regulator of immune exhaustion in other biological contexts.^9^ In preclinical animal models of H1N1 influenza and lymphocytic choriomeningitis virus infection, anti-TNFα monotherapy resolved inflammation, restored immune surveillance, suppressed or cleared viral infection and prolonged survival.^9,10^ Based on the hypothesis that TNFα-antagonists may abrogate the adverse inflammatory cytokine profile of COVID-19 disease and reduce the need for advanced cardiorespiratory support, a pilot study of the infliximab biosimilar, infliximab-abda, was initiated in COVID-19 respiratory failure (NCT04425538). The goal of the study was to generate early efficacy, toxicity and cytokine correlative data to inform the next generation of studies.

## Methods

### Patients (Figure 1)

Eligible subjects were at least 18 years old, could provide informed consent, had pneumonia evidenced by chest X-ray or computerized tomography, and laboratory (reverse transcriptase-polymerase chain reaction) confirmed infection for severe acute respiratory syndrome coronavirus 2 (SARS-CoV-2) with at least one of the following: respiratory frequency ≥30/min, blood oxygen saturation ≤93% on room air, partial pressure of arterial oxygen to fraction of inspired oxygen ratio (PaO2/FiO2) <300, worsening of lung involvement defined as an increase in number and/or extension of pulmonary areas of consolidation, need for increased FiO2 to maintain stable oxygen saturation, or worsening oxygen saturation of >3% with stable FiO2. Exclusion criteria included treatment with any TNFα inhibitor in the past 30 days, absolute neutrophil count less than 1000 mm3, hemoglobin <7.0g/L, platelets <50,000 per mm3, or aspartate transaminase or alanine transaminase greater than 5 times the upper limit of normal, known active or latent hepatitis B, known or suspected active tuberculosis (TB) or a history of incompletely treated or latent TB, pregnancy, uncontrolled systemic bacterial or fungal infections (prior positive bacterial or fungal cultures on appropriate therapy with negative repeat cultures was permissible), myocardial infarction (within last month), moderate or severe heart failure (New York Heart Association class III or IV), acute stroke (within last month), uncontrolled malignancy, Stage 4 severe chronic kidney disease or requiring dialysis (i.e. estimated glomerular filtration rate (eGFR) < 30 ml /min/1.73 m2) at baseline. Prior and concurrent remdesivir, dexamethasone and convalescent plasma therapy were permitted. No limits to the level and duration of oxygenation support or hospitalization were specified, Infliximab-abda therapy was sanctioned as part of a COVID-19 care pathway for hospitalized patients by an institutional task force and by infectious disease and critical care medicine consultation for each patient. Patients who were judged to be improving on standard therapy or suspected to have active concomitant infection were not sanctioned for study consideration.

**Figure 1.**
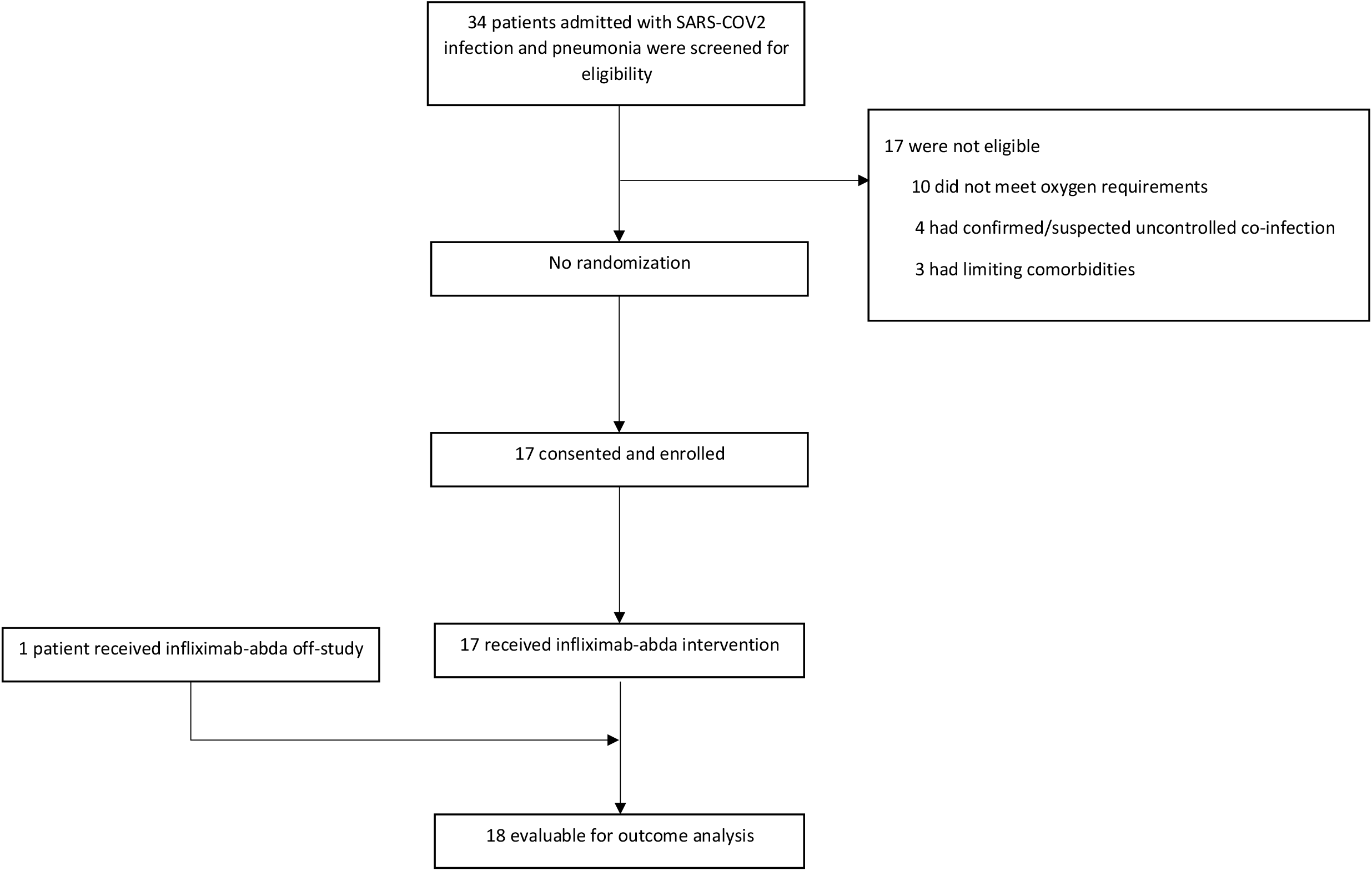
Consort diagram. Hospitalized patients with SARS-COV2 infection and pneumonia that were referred to the infliximab-abda study team for evaluation.

### Therapy

Treatment with infliximab-abda 5mg/kg IV was planned within 6 hours of enrollment, and no more than 24 hours following enrollment. Pre-medication with acetaminophen 650 mg once, 30 minutes prior to infusion, was recommended. Retreatment with infliximab-abda was permitted at the treating physician’s discretion 7-21 days following primary therapy. One patient (TMC-Pre) who met the eligibility criteria and was treated on the care pathway prior to the formal activation of the study was included in the analysis and reporting.

### Laboratory studies

included complete blood counts and leucocyte differential, complete metabolic panels which included electrolytes, creatinine and liver function tests, inflammatory markers including serum ferritin and C-reactive protein (CRP) and a panel of cytokines performed at baseline prior to infliximab-abda (day 1) in all enrolled patients and on Days 3 and 14 or discharge if earlier. Luminex xMAP technology was employed for multiplexed quantification of 48 human cytokines, chemokines, and growth factors. The multiplexing analysis was performed using the Luminex™ 200 system (Luminex, Austin, TX, USA) by Eve Technologies Corp. (Calgary, Alberta). Forty-eight markers were simultaneously measured in the samples using Eve Technologies’ Human Cytokine 48-Plex Discovery Assay® (MilliporeSigma, Burlington, Massachusetts, USA) according to the manufacturer’s protocol. The 48-plex consisted of sCD40L, EGF, Eotaxin, FGF-2, FLT-3 Ligand, Fractalkine, G-CSF, GM-CSF, GROα, IFN-α2, IFN-γ, IL-1α, IL-1β, IL-1RA, IL-2, IL-3, IL-4, IL-5, IL-6, IL-7, IL-8, IL-9, IL-10, IL-12 (p40), IL-12(p70), IL-13, IL-15, IL-17A, IL-17E/IL-25, IL-17F, IL-18, IL-22, IL-27, IP-10, MCP-1, MCP-3, M-CSF, MDC, MIG/CXCL9, MIP-1α, MIP-1β, PDGF-AA, PDGF-AB/BB, RANTES, TGFα, TNF-α, TNF-β, and VEGF-A. Assay sensitivities of these markers range from 0.14 – 50.78 pg/mL.

### Endpoints

The primary endpoint of the pilot study was time to improvement in oxygenation (increase in oxygen saturation to fraction of inspired oxygen ratio SpO2/FiO2 of 50 or greater compared to the baseline SpO2/FiO2) sustained for a minimum of 48 hours. Secondary endpoints included 28-day mortality, assessment of dynamic changes in cytokine and inflammatory profile after therapy, qualitative and quantitative toxicity, incidence and duration of supplementation oxygen support including mechanical ventilation and extracorporeal membrane oxygenation (ECMO), duration of hospitalization and frequency and spectrum of secondary infections. COVID-19-associated pulmonary aspergillosis diagnoses were annotated by consensus criteria.^11^

### Statistics

Mean ferritin, CRP, and cytokine levels on Day 1 and the mean dynamic change on Day 3 and Day 14 were compared using pairwise ratio t-tests (GraphPad Prism). Given the number of comparisons (50 at each timepoint), a 5% false-discovery rate (FDR) was applied using the Benjamini-Hochberg method, and those cytokines that met statistical significance at the adjusted thresholds are also reported. Given that post-steroid therapy IL-6 levels at different cut-off levels have been shown to correlate with adverse outcomes^12,13^, the impact of infliximab-abda on IL-6 levels at Day 3 and Day 14 stratified by degree of baseline elevation in the largely steroid-pretreated population was separately analyzed. Pearson correlation coefficients were calculated for fold changes (log2 transformed ratio) in expression from baseline to day 3 between each pair of cytokines. In all statistical comparisons, values for samples whose cytokine levels were below the assay detection limits were replaced by the sensitivity of the assay for each respective cytokine. Descriptive statistics annotate the remaining endpoints.

### Study approval

The study was approved by the institutional review board and written informed consent was received from participants or designees prior to inclusion.

## Results

The demographics, clinical features, inflammatory markers and outcomes of 18 patients with COVID-19 respiratory failure treated with infliximab-abda between April and December 2020 are represented in Table 1. The median age was 63 years (range 31-80), largely male (78%), and race-ethnic minority (Asian, South Asian, Black, Hispanic or Latino, 72%) with at least one significant medical comorbidity (72%). Comorbidities included hypertension (67%), diabetes (28%), smoking history (22%) or chronic lung disease, including asthma (22%). Half were obese or morbidly obese; the median body mass index was 30.3 kg/m2 (range 18.0-41.8). The median Charlson Comorbidity Index was 2.5 (range 0-9). Sixteen patients (89%) had received both remdesivir and steroid therapy (median 2 days, range 0-7 days) with infliximab-abda therapy supported by the infectious disease consultative service. Five patients (28%) received prior convalescent plasma (median 2 days, range 0-4 days). Fifteen patients (83%) were on high-flow nasal cannula (HFNC), mechanical ventilation (MV) and/or extracorporeal membrane oxygenation (ECMO) and four (22%) were receiving vasopressor support. The median ferritin level was 1953 ng/ml and 13/18 (72%) of patients were lymphopenic. The median baseline SpO2/FiO2 was 202 (range 110-336). The median duration from symptom onset to infliximab-abda therapy was 10 days (range, 5-29 days), and the median duration from hospitalization to infliximab-abda therapy was 2 days (range, 0-7 days).

|  | **TMC-**<br>**OFF** | **TMC-**<br>**001** | **TMC-**<br>**002** | **TMC-**<br>**003** | **TMC-**<br>**004** | **TMC-**<br>**005** | **TMC-**<br>**006** | **TMC-**<br>**007** | **TMC-**<br>**008** | **TMC-**<br>**009** | **TMC-**<br>**010** | **TMC-**<br>**011** | **TMC-**<br>**012** | **TMC-**<br>**013** | **TMC-**<br>**014** | **TMC-**<br>**015** | **TMC-**<br>**016** | **TMC-**<br>**017** |
| --- | --- | --- | --- | --- | --- | --- | --- | --- | --- | --- | --- | --- | --- | --- | --- | --- | --- | --- |
| **Age (range, years)** | >60 | >60 | 21-40 | >60 | 41-60 | >60 | 41-60 | >60 | 41-60 | >60 | >60 | 41-60 | >60 | >60 | 41-60 | 41-60 | >60 | 41-60 |
| **Sex** | Female | Male | Male | Male | Male | Male | Female | Male | Male | Male | Male | Female | Male | Male | Female | Male | Male | Male |
| **Race or ethnic group minority (Yes/No)** | Yes | Yes | No | Yes | Yes | No | Yes | No | Yes | Yes | Yes | No | Yes | No | Yes | Yes | No | Yes |
| **Charlson Comorbidity Index** | 6 | 6 | 0 | 4 | 1 | 3 | 0 | 3 | 1 | 2 | 3 | 1 | 9 | 4 | 2 | 2 | 5 | 2 |
| **Baseline Laboratory Values** |  |  |  |  |  |  |  |  |  |  |  |  |  |  |  |  |  |  |
| Lymphocyte Count (K/uL) | 0.8 | 0.8 | 1.4 | 0.7 | 1.3 | 0.2 | 0.4 | 0.4 | 0.5 | 0.9 | 0.7 | 0.8 | 1 | 1.2 | 0.4 | 0.5 | 0.7 | 1 |
| LDH (IU/L) | 503 | 372 | 544 | 403 | 375 | 313 | 431 | 493 | 415 | 321 | 613 | 490 | 595 | 835 | 313 | 361 | 599 | 339 |
| Ferritin (ng/mL) | 4773 | 1407 | 2244 | 1598 | 1273 | 2013 | 352 | 2128 | 343 | 6279 | 2063 | 2008 | 2782 | 1898 | 221 | 2369 | 1122 | 626 |
| D-dimer (ng/mL) | 1969 | 710 | 264 | 420 | 183 | 353 | 343 | 267 | 491 | <150 | 485 | 998 | 2199 | 531 | 176 | 194 | 843 | <150 |
| CRP (mg/L) | 33.6 | 243.2 | 64.1 | 200.1 | 54.4 | 108.6 | 233.6 | 104.3 | 58.4 | 57.9 | 164.4 | 54.3 | 140.3 | 29.7 | 214.3 | 26.9 | 50.7 | 7.87 |
| **Vasopressor support** | Yes | No | No | Yes | No | Yes | No | No | No | No | No | No | No | Yes | No | No | No | No |
| **Baseline SpO2/FiO2** | 230 | 188 | 332 | 111 | 214 | 145 | 261 | 209 | 196 | 182 | 134 | 271 | 110 | 93 | 336 | 220 | 243 | 139 |
| **Oxygenation Support\*** |  |  |  |  |  |  |  |  |  |  |  |  |  |  |  |  |  |  |
| Time of enrollment | MV | HFNC | NC | MV | NC | MV | HFNC | HFNC | MV | HFNC | MV | HFNC | ECMO | MV | NC | HFNC | HFNC | HFNC |
| Maximum support | MV | HFNC | NC | MV | NC | MV | HFNC | HFNC | MV | HFNC | MV | HFNC | ECMO | MV | NC | MV | MV | HFNC |
| Discharge or death | NC | RA | RA | RA | RA | RA† | RA | NC | RA | NC | MV | NC | RA† | MV | RA | RA† | MV | RA |
| **Symptom Onset to infliximab-abda (days)** | 29 | 15 | 12 | 13 | 5 | 27 | 5 | 9 | 10 | 10 | 7 | 8 | 10 | 12 | 11 | 8 | 20 | 12 |
| **Admission to Infliximab-abda (days)** | 6 | 3 | 5 | 1 | 3 | 7 | 2 | 2 | 2 | 2 | 0 | 1 | 2 | 3 | 1 | 2 | 1 | 5 |
| **Hospital duration following infliximab-abda (days)** | 21 | 8 | 1 | 14 | 5 | 47 | 7 | 12 | 12 | 7 | 22 | 4 | 52 | 10 | 4 | 37 | 34 | 9 |
| **Primary Endpoint Met (Day met)** | Yes (12) | Yes (3) | Yes (1) | Yes (3) | Yes (3) | Yes (3) | Yes (6) | No | Yes (6) | Yes (3) | Yes (6) | Yes (5) | Yes (4) | Yes (2) | Yes (4) | No | Yes (12) | Yes (4) |
| **Secondary infections** | Yes | No | No | Yes | No | Yes | No | No | No | No | Yes | No | Yes | Yes | No | Yes | Yes | No |
| **Survival to discharge** | No | Yes | Yes | Yes | Yes | Yes | Yes | Yes | Yes | Yes | No | Yes | Yes | No | Yes | Yes‡ | No | Yes |

\*Within 24 hours of trial enrollment; †Via tracheostomy; ‡Ventilated at day 28
RA- Room air; NC- Nasal Cannula; HFNC- High Flow Nasal Cannula; MV- mechanical ventilator; ECMO- Extracorporeal membrane oxygenation

Following infliximab-abda therapy, sixteen patients (89%) met the primary endpoint of sustained improvement in SpO2/FiO2 of ≥50 for at least 48 hours and fifteen patients (83%) had resolution of respiratory failure to room air or nasal cannula (NC) oxygen supplementation. The median time to the primary endpoint was 4 days (range, 1-12 days). The median support duration among patients requiring NC was 3 days (range, 1-4 days), for HFNC 2.5 days (range, 2-6 days), and MV 10 days (range, 5-45 days) (Figure 2). One patient on ECMO was successfully de-cannulated after 16 days. One patient on MV with prior steroid and tocilizumab-failure and on multiple vasopressors was successfully extubated to full recovery of respiratory failure on room air. No patient received a second dose of infliximab-abda. Fifteen patients (83%) were alive at the end of the 28-day study period, with one death occurring on day 34. Fourteen patients (78%) were discharged after a median of 8 days (range 1-52 days), 3 on oxygen supplementation via NC and 11 on room air.

**Figure 2.**
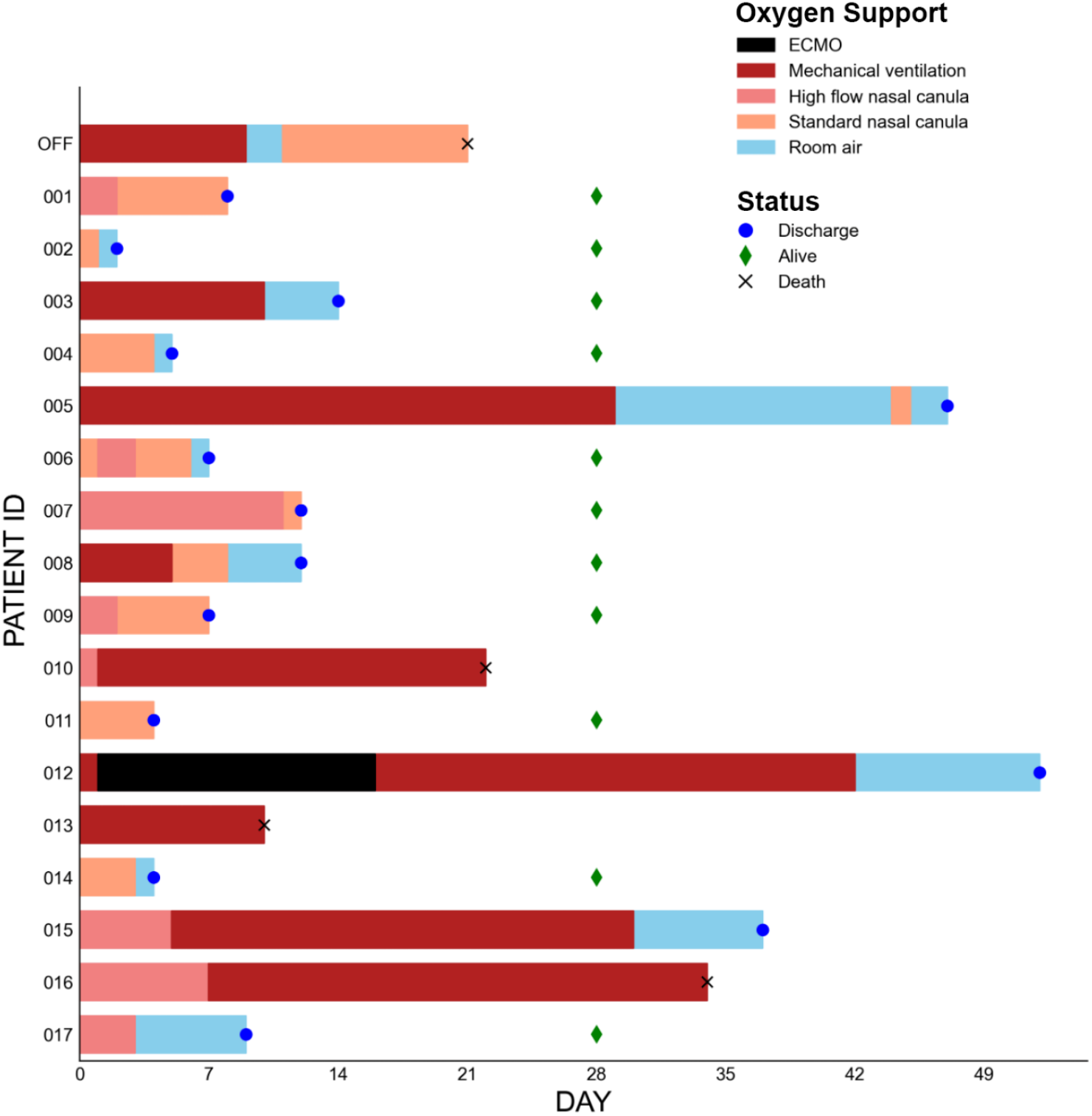
Changes in oxygen support status following infliximab-abda treatment. Colored bars indicate the maximal level of oxygen support for each individual following treatment with infliximab-abda. The status of the patient at last follow-up (discharged, alive or dead) is indicated. ECMO: extracorporeal membrane oxygenation

By Day 3 there was a significant decline from elevated baseline levels in a subset of pro-inflammatory and immunomodulatory cytokines which have been implicated in pathogenesis and/or adverse outcomes from severe COVID-19 illness, including IP-10, IL-10, IL-27, TNF-α and IFN-γ ^1,5,8,14-16^ (Figure 3, Supplemental Table 1). Among patients with elevated IL-6 levels at baseline (≥ 10pg/ml), including one without steroid exposure, sharp declines at Day 3 were uniformly observed (Figure 3, Supplemental Figures 1-2). CRP and ferritin were significantly lower as well. Reduction of the CXCR3-ligands IP-10 and CXCL-9,^8,14^ IL-27, CRP and ferritin at Day 14 were sustained although only IP-10 stood up to adjustment for multiple comparisons among the 50 cytokines and markers assessed. The majority of inflammatory and immunomodulatory cytokines assessed in the 48-plex assay did not change significantly, either at Day 3 or Day 14 (Figure 3, Supplemental Table 1).

**Figure 3.**
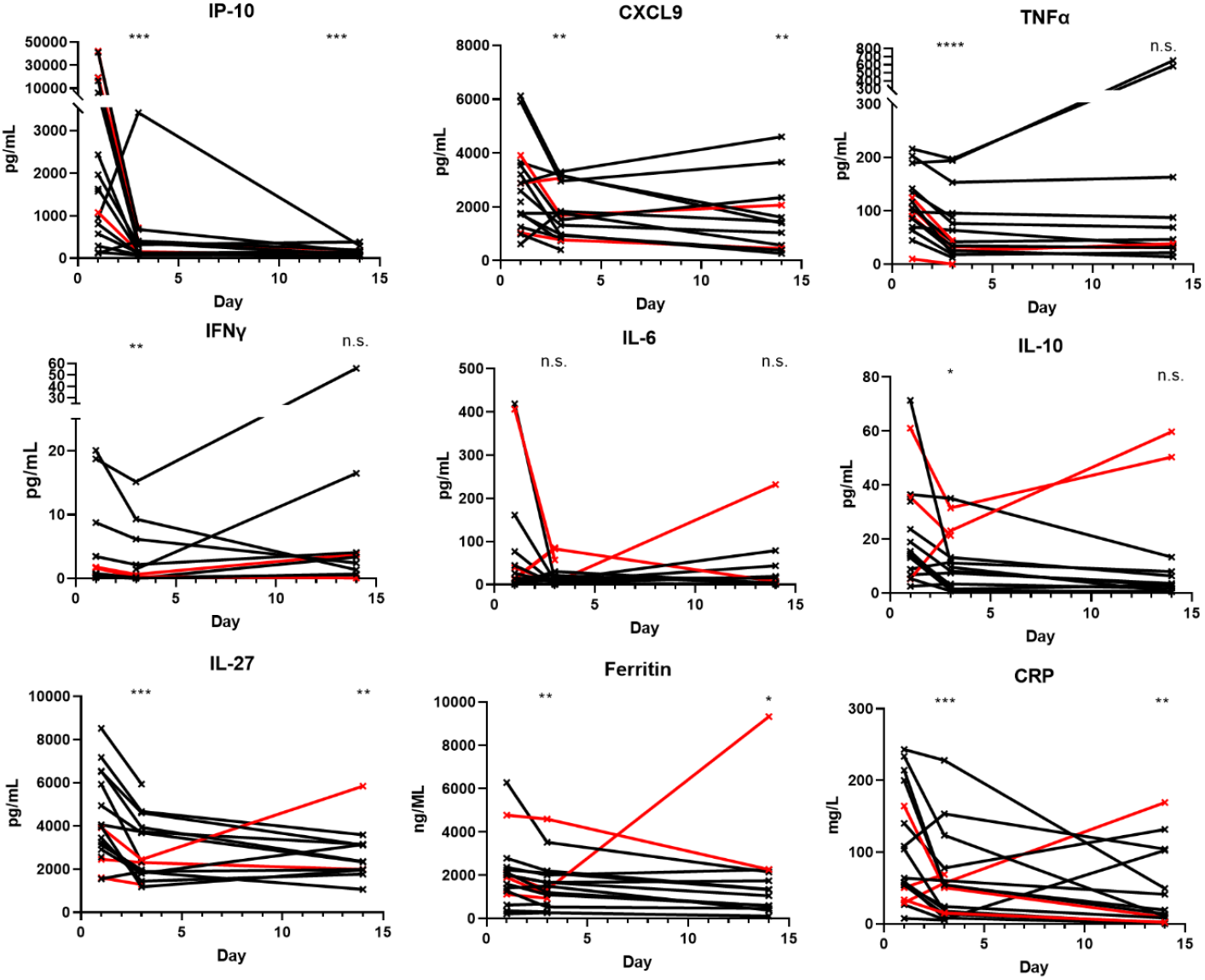
Decline in key cytokines and inflammatory markers following infliximab-abda therapy. Values from individuals are connected with solid lines, with deceased individuals indicated in red. Statistics: n=18, paired ratio t-test compared to baseline; *: P< 0.05, **: P<0.01, ***: P<0.001, ****: P<0.0001, n.s.: not significant.

Among 13 patients who were lymphopenic at baseline, six (46%) had complete resolution of lymphopenia by day 3, and 11 recovered by day 14. Absolute lymphocyte counts (normal 1.0-4.0 K/µl) increased from an average baseline of 0.8 to 1.0 K/µl by day 3 (P=0.038, n=17) and 1.7 K/µl by day 14/discharge (P=0.0004, n=13). Absolute monocyte counts (normal 0.2-0.8 K/µl) also increased from an average baseline of 0.5 to 0.8 K/µl by day 3 (P= 0.0045, n=17) and 1.1 K/µl by day 14/discharge (P=0.002, n=13). Accordingly, the strongest correlation between dynamic changes in IP-10 among all tested cytokines between baseline and Day 3 (Supplemental Table 2) was with CXCL-9 (Pearson r: 0.653, p-value: 0.004, n=17), in particular among lymphopenic patients (Pearson r: 0.843, p-value: 0.0003, n=13) and those that recovered by Day 3 (Pearson r: 0.98, p-value: 0.0006, n=6) and Day 14 (Pearson r: 0.85, p value 0.008, n=8 evaluable), whereas this correlation was not detectable among the small number of non-lymphopenic patients (Pearson r: -0.217, p-value: 0.782, n=4). Among the non-lymphopenic patients, IP-10 decline correlated instead with IL12p40, a homodimeric inhibitory subunit of IL-12.

Infliximab-abda was well tolerated. Two patients experienced transient Grade 1 infusion-related reactions. Transaminase elevations occurred in 11 patients but association with infliximab-abda could not be inferred. There were no cases of deep venous thrombosis or pulmonary thromboembolism. Transient clinical worsening among 2 patients following rapid lymphocyte recovery raised suspicions of an immune reconstitution inflammatory syndrome (IRIS). One patient with suspected IRIS received high-dose steroids with subsequent clinical improvement but later succumbed to secondary infection.

All active infections were considered as controlled at the time of infliximab-abda administration. Seventeen secondary infections were encountered in eight patients (44%), all of whom were mechanically ventilated or on ECMO for critical respiratory failure. Six were lymphopenic, four patients were diabetic, three had chronic lung disease, seven had hypertension, and all had prior steroid therapy for a median duration of five days. The majority of enrolled patients with diabetes (4/5), chronic lung disease (3/4), hypertension (7/12) and those on MV/ECMO support (8/9) were eventually diagnosed with secondary infections. Nine infections (53%) were bacterial, four (24%) were fungal, and four (24%) were viral in etiology (Supplemental Table 3). Median time to diagnosis was 9 days (range, 1-35 days). The most common diagnosis was ventilator-associated pneumonia (41%). Two instances of herpes simplex virus pneumonitis were observed, one of which was diagnosed at autopsy with concomitant necrotizing pyogenic infection in a patient with underlying diabetes, hypertension and obesity who had received high-dose parenteral steroids. One possible and two probable instances of COVID-19-associated pulmonary aspergillosis (CAPA) were encountered, of whom 2 died. Both deaths occurred in patients with pre-existing chronic lung disease. Two patients (11%) were diagnosed with low-level cytomegalovirus viremia, not requiring treatment. One death in a frail elderly patient was attributed to aspiration and upper gastrointestinal hemorrhage following recovery to room air oxygenation after prolonged mechanical ventilation. The deaths on study did not correlate with the improvements with oxygenation as specified in the primary endpoint (Table 1), but with late-emerging events during hospitalization as described.

## Discussion

Notwithstanding the uncontrolled nature of these observations, the clinical and translational outcomes following infliximab-abda administration are consistent with the hypothesis that TNFα is a major regulator of the adverse cytokine signature and pathobiology of severe and critical COVID-19 respiratory failure. One of the most striking outcomes was the rapid and sustained decline in the markedly elevated levels of the T-cell chemokine IP-10, which along with CXCL-9, is potentially generated by aberrantly amplified synergy between IFN-γ and TNFα as previously proposed from preclinical modeling^7^, and which has strongly correlated with adverse outcomes.^8^ IP-10 has been mechanistically implicated in the pathogenesis of adult respiratory distress syndrome (ARDS)^17^ and elevations of IP-10 in COVID-19 may predict the duration of mechanical ventilation.^18^ Our pilot data integrated with the mechanistic studies from preclinical models, predict that TNFα-antagonist therapy, specifically infliximab-abda, may rapidly control IP-10 and associated pathological inflammatory signaling. Rapid resolution of lymphopenia in our study may reflect the coordinate drop in IP-10 and CXCL-9 which drive tissue redistribution of activated and/or exhausted T-lymphocytes via CXCR3 receptors.^7,19^ Upregulation of CXCR3 ligands in the lung microenvironment may similarly drive peripheral depletion of CXCR3+ monocytes with induction of macrophage differentiation, which additionally sustain and amplify the inflammatory loop.^20-22^ Increasing monocyte counts following infliximab-abda therapy may also reflect reversal of this process.

The study was conducted during a time in which the standards of care among hospitalized patients evolved to integrate remdesivir and dexamethasone, which nearly all patients received prior to infliximab-abda administration. Although clinicopathological outcomes arguably reflect the combination of these agents, in addition to convalescent plasma and supportive antimicrobial therapies, patients improving on standard therapy were not sanctioned for the study by infectious disease consultation. Furthermore, one patient with rapid clinical response prior to discharge did not receive prior remdesivir or glucocorticoid therapy. The cytokine profile following infliximab-abda therapy in this patient exemplifies the rapid and sustained reductions from markedly elevated baseline values of IP-10, IL-6, IL-10, CXCL-9 and TNFα (Supplemental Figure 2) seen in the majority of patients, demonstrating that the clinicopathological outcome can be independent of steroid therapy. In vitro models demonstrated the lack of impact of glucocorticoids on IP-10 and CXCL-9 secretion secondary to IFNγ and TNFα.^7^ This suggests that TNFα-blockade may represent a therapeutic strategy that could offer more precise and durable control of the hyperinflammatory cytokine signature of COVID-19 than the broad-spectrum anti-inflammatory effects of steroid therapy. Furthermore, IL-6 could not induce IP-10 and CXCL-9 secretion ^7^ which may also infer a limit to the effectiveness of IL-6-directed therapies in controlling the broader hyperinflammatory picture of serious COVID-19 illness.

Importantly, serial cytokine profiling from steroid and IL-6-directed randomized clinical trials have not been reported to allow formal assessment of these points.^23-25^ The high rate of clinical recovery and the cytokine profiling suggests that dual TNFα and IFN-γ targeting, which has been proposed based on preclinical modeling of lethal COVID-19, ^1^ may not be necessary. The attenuated yet elevated levels of CXCL-9 contrasted with uniformly diminished IP-10 levels at Day 14 may reflect restoration of physiological IFN-γ-mediated immunosurveillance, rather than persistent pathological signaling.^7,19^

In contrast to the lymphopenic subgroup, a strong association between declines in IP-10 and IL12p40 (and not IL12p70) was suggested among the non-lymphopenic patients. IL-12 directs proliferation of activated T lymphocytes towards a Th1 phenotype. The heterodimeric molecule IL-12p70, equates with IL-12 biological activity, whereas IL-12p40 may antagonize IL-12 and inhibit cytotoxic T lymphocyte generation^26^. Increased IL-12 levels derived from macrophages and dendritic cells contribute to the heterogeneity of cytokine storms^27^ and restoration of physiological IL12p40:IL-12p70 ratios may recover T-cell function.

Another noteworthy finding was the early and sustained decline in IL-27 (Figure 2) which has been co-implicated with TNFα in association with disease severity among older patients (>60 yrs), but in a cytokine network distinct from TNFα.^16^ IL-27 may drive an IFN-γ/TNFα-independent program of immune exhaustion, via potent induction of Tim-3 and IL-10 ^28^. In keeping with this potential mechanism of treatment resistance to TNFα-antagonists, two of four patients who died were evaluable with cytokine assays at Day 14; both had rising levels of IL-10, one with a rising IL-27 (Figure 3).

The frequency and spectrum of secondary infections observed in this study are readily comparable to those observed among a mechanically ventilated population with COVID-19 respiratory failure with similar indices of disease severity.^29,30^ Nevertheless, this requires additional scrutiny in randomized studies of TNF-antagonists, with particular caution among lymphopenic patients on mechanical ventilation with pre-existing lung disease and/or multiple comorbid factors such as diabetes or prolonged/high-doses of steroids.^31,32^ Herpes simplex reactivation has been associated with prolonged mechanical ventilation and may be routinely underdiagnosed.^30,33^ Similarly, estimates of CAPA in COVID-19 has varied widely, up to 35% of patients with ARDS.^31^ Unfortunately, autopsy permission was granted in only one of three infection-related deaths on study.

A placebo-controlled randomized clinical trial conducted by the National Institutes of Health (ACTIV-1, NCT04593940) features eligibility criteria and a treatment plan similar to this study, integrating an infliximab arm combined with remdesivir, and permitting dexamethasone and convalescent plasma. The principal endpoint of ACTIV-1 is an improvement in time to recovery within 29 days with mortality as a secondary endpoint. Given that dexamethasone is likely to be widely employed in both control and experimental arms, whether TNFα-antagonist monotherapy will offer greater precision, clinical efficacy and safety over dexamethasone or other cytokine-directed therapeutics in severe and critical COVID-19 respiratory failure may remain an open question. While serial cytokine assays tethered to cellular markers of immune exhaustion will shed additional light into disease heterogeneity, response and resistance to TNFα-antagonists, the importance of survival as the optimal principal endpoint of adequately powered trials has been emphasized.^25^

## Data Availability

Clinical and cytokine data are represented in the report and may be supplemented e.g. raw cytokine data, on request.

## Author contributions

Hilal Hachem, supported trial design, grant application and study accrual; Amandeep Godara, supported trial design, grant application, study accrual and set up translational cytokine assays, Courtney Schroeder, supported trial design, study accrual, data analysis and reporting, Daniel Fein, supported study accrual, data analysis and reporting, Hashim Mann supported study accrual, data analysis and reporting, Christian Lawlor supported trial design, study accrual, managed translational specimens and data quality, Jill Marshall supported study accrual, processing of translational specimens, and data management, Andreas Klein supported trial design, grant application and study reporting, Debra Poutsiaka supported trial design, grant application and study reporting, Janis Breeze assisted with statistical design and reporting, Raghav Joshi supported data analysis and study reporting, Paul Mathew conceived and designed the study, prepared the grant application, managed the study group and reported the results. There were no conflicts of interest identified among all authors.

## Acknowledgements

Supported by the National Center for Advancing Translational Sciences, National Institutes of Health, Award Number UL1TR002544. The content is solely the responsibility of the authors and does not necessarily represent the official views of the NIH. The administrative support of Ms Vidya Iyer and the multidisciplinary clinical teams involved in the care of patients on the study is acknowledged.

