## Supplemental Data for "Rapid and sustained decline in CXCL-10 (IP-10) annotates clinical outcomes following TNF-α antagonist therapy in hospitalized patients with severe and critical COVID-19 respiratory failure"

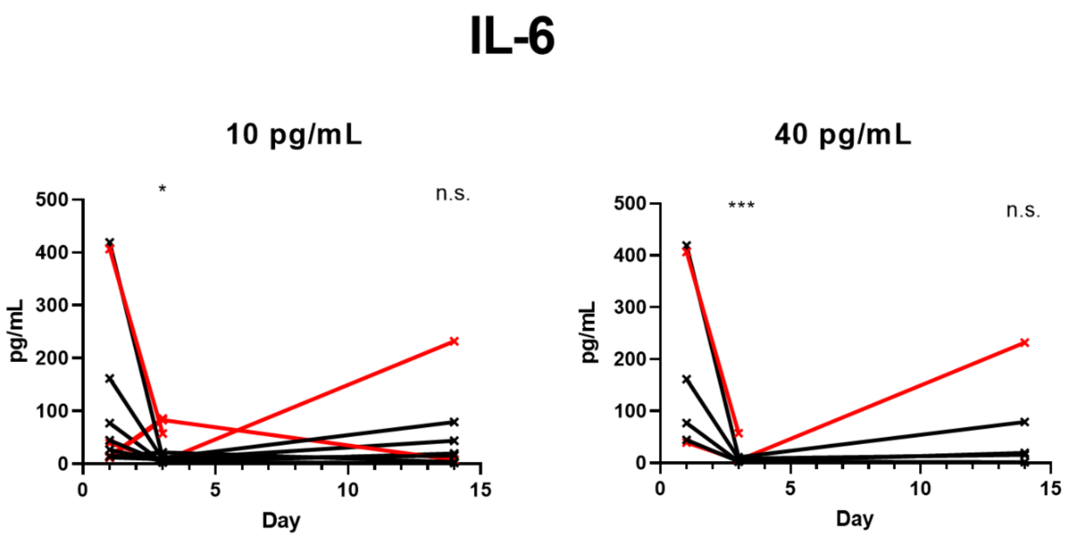


**Figure 1. Control of IL-6 in patients stratified by variable baseline elevations.** Inflammatory marker and cytokine expression was assessed by multiplex ELISA at days 0, 3 and 14 (or discharge). Expression values from individuals are connected with solid lines, with deceased individuals indicated in red. Values are plotted with exclusion of patients with baseline IL-6 expression lower than 10 (n=11) and 40 (n=6) pg/mL. Statistics: Paired ratio t-tests were used to compare the expression levels at days 3 and 14 to baseline. *: P< 0.05, ***: P<0.001, n.s.: not significant.

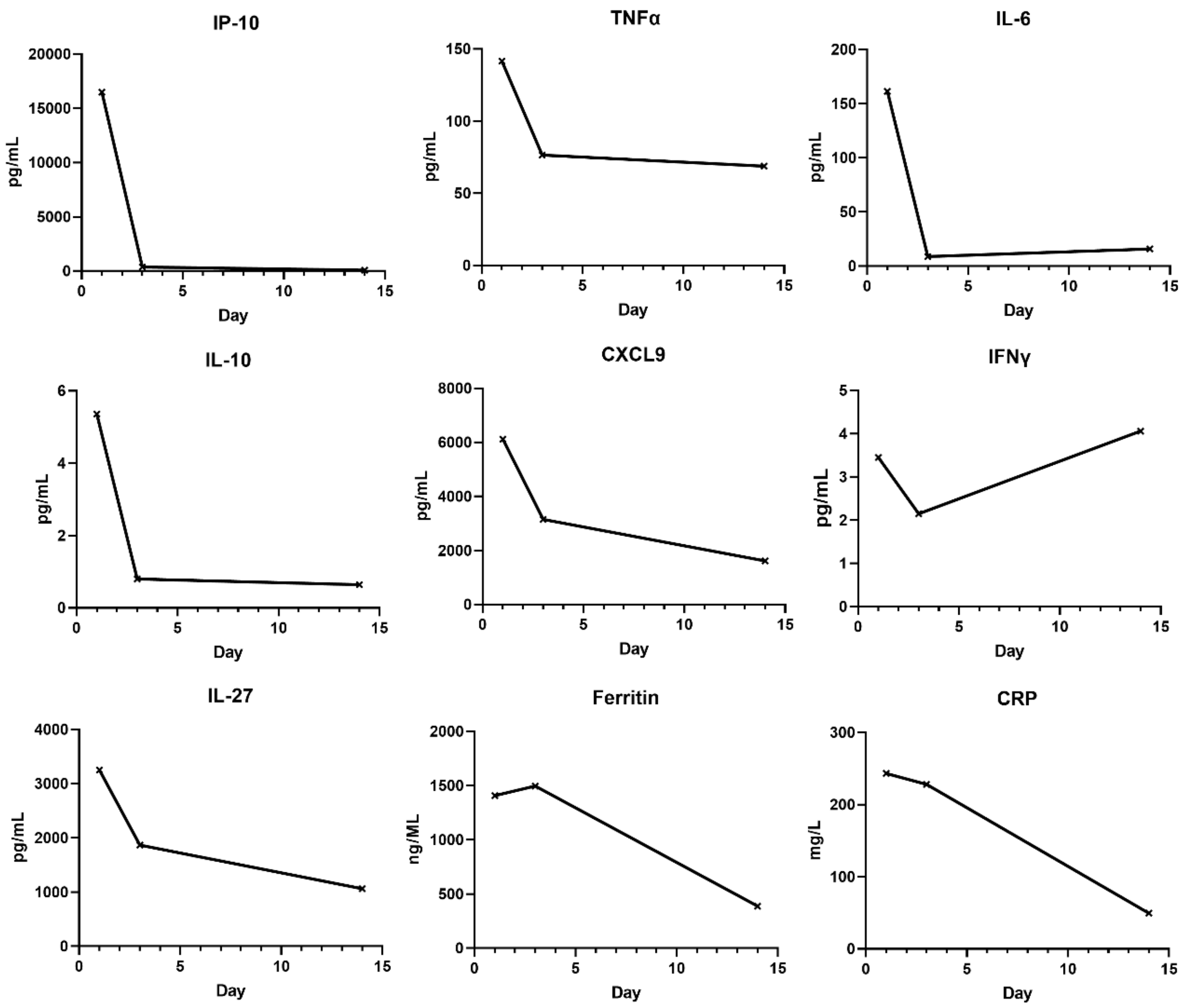


**Figure 2. Control of inflammatory markers and cytokines following infliximab therapy in a patient without prior steroid or remdesivir exposure.**

**Table 1. Change in cytokine levels at Days 3 and 14**

| Day 3 | | | Day 14 | | |
|---|---|---|---|---|---|
| **Cytokine** | **P-value** | **FDR Critical value** | **Cytokine** | **P-value** | **FDR Critical value** |
| TNFa | 0.000071 | 0.001 | IP-10 | 0.00055 | 0.001 |
| IL-27 | 0.00012 | 0.002 | CRP | 0.003039 | 0.002 |
| IP-10 | 0.00016 | 0.003 | IL-27 | 0.0088 | 0.003 |
| CRP | 0.00058 | 0.004 | CXCL9 | 0.0088 | 0.004 |
| IFNy | 0.002 | 0.005 | Ferritin | 0.013907 | 0.005 |
| Ferritin | 0.002098 | 0.006 | IL-22 | 0.017 | 0.006 |
| CXCL9 | 0.0088 | 0.007 | IL-5 | 0.021 | 0.007 |
| IL-12p40 | 0.011 | 0.008 | GROa | 0.026 | 0.008 |
| IL-10 | 0.013 | 0.009 | TNFa | 0.056 | 0.009 |
| Eotaxin | 0.035 | 0.01 | VEGF-A | 0.064 | 0.01 |
| IL-4 | 0.067 | 0.011 | IL-10 | 0.072 | 0.011 |
| IL-6 | 0.069 | 0.012 | IL-3 | 0.073 | 0.012 |
| GM-CSF | 0.081 | 0.013 | G-CSF | 0.15 | 0.013 |
| MCP-1 | 0.089 | 0.014 | IL-7 | 0.2 | 0.014 |
| MDC | 0.097 | 0.015 | Fractalkine | 0.22 | 0.015 |
| MCP-3 | 0.097 | 0.016 | FLT-3L | 0.22 | 0.016 |
| M-CSF | 0.098 | 0.017 | IL-17E/IL-25 | 0.22 | 0.017 |
| IL-8 | 0.14 | 0.018 | IL-6 | 0.24 | 0.018 |
| IL-17A | 0.16 | 0.019 | IL-15 | 0.26 | 0.019 |
| Fractalkine | 0.16 | 0.02 | IL-17F | 0.26 | 0.02 |
| EGF | 0.17 | 0.021 | GM-CSF | 0.27 | 0.021 |
| IL-13 | 0.17 | 0.022 | IL-4 | 0.28 | 0.022 |
| VEGF-A | 0.18 | 0.023 | FGF-2 | 0.29 | 0.023 |
| GROa | 0.21 | 0.024 | Eotaxin | 0.33 | 0.024 |
| FLT-3L | 0.22 | 0.025 | EGF | 0.33 | 0.025 |
| sCD4OL | 0.26 | 0.026 | IL-18 | 0.33 | 0.026 |
| G-CSF | 0.3 | 0.027 | IL-1RA | 0.33 | 0.027 |
| IL-9 | 0.38 | 0.028 | TNFB | 0.38 | 0.028 |
| IL-12p70 | 0.39 | 0.029 | MCP-3 | 0.39 | 0.029 |
| IL-18 | 0.41 | 0.03 | IL-12p40 | 0.4 | 0.03 |
| TGFa | 0.41 | 0.031 | IL-9 | 0.41 | 0.031 |
| IL-la | 0.42 | 0.032 | IFN-a2 | 0.46 | 0.032 |
| FGF-2 | 0.45 | 0.033 | RANTES | 0.48 | 0.033 |
| TNFB | 0.45 | 0.034 | IL-1B | 0.5 | 0.034 |
| IL-22 | 0.46 | 0.035 | sCD4OL | 0.53 | 0.035 |
| IFN-a2 | 0.49 | 0.036 | IL-8 | 0.55 | 0.036 |
| IL-2 | 0.52 | 0.037 | IL-2 | 0.65 | 0.037 |
| IL-1B | 0.66 | 0.038 | TGFa | 0.66 | 0.038 |
| IL-17E/IL-25 | 0.66 | 0.039 | MIP-la | 0.67 | 0.039 |
| IL-15 | 0.68 | 0.04 | PDGF-AB/BB | 0.68 | 0.04 |
| IL-5 | 0.7 | 0.041 | IL-13 | 0.69 | 0.041 |
| MIP-1B | 0.7 | 0.042 | MIP-1B | 0.73 | 0.042 |
| PDGF-AB/BB | 0.71 | 0.043 | MDC | 0.74 | 0.043 |
| IL-17F | 0.88 | 0.044 | IL-17A | 0.77 | 0.044 |
| PDGF-AA | 0.89 | 0.045 | M-CSF | 0.78 | 0.045 |
| IL-7 | 0.9 | 0.046 | MCP-1 | 0.82 | 0.046 |
| IL-3 | 0.92 | 0.047 | IL-12p70 | 0.83 | 0.047 |
| RANTES | 0.93 | 0.048 | IL-la | 0.84 | 0.048 |
| IL-1RA | 0.94 | 0.049 | IFNy | 0.87 | 0.049 |
| MIP-la | 0.98 | 0.05 | PDGF-AA | 0.98 | 0.05 |

A 5% false-discovery rate (FDR) adjustment was applied using the Benjamini-Hochberg method. The cytokines are ordered in ascending p-value; green highlighting denotes statistical significance after the FDR correction.

### Table 2: Pearson correlation between dynamic changes in IP-10 with other cytokines

| Total (n=17) | | Lymphopenic (n=13) | | Recovered* by day 3 (n=6) | | Non-lymphopenic (n=4) | |
|---|---|---|---|---|---|---|---|
| **MIG/CXCL9** | **0.65** | **MIG/CXCL9** | **0.84** | **MIG/CXCL9** | **0.98** | IL-12p40 | 0.99 |
| IL-15 | 0.60 | IL-3 | 0.71 | M-CSF | 0.83 | sCD40L | 0.95 |
| FLT-3L | 0.60 | IL-15 | 0.70 | IL-1RA | 0.74 | EGF | 0.93 |
| IL-12p40 | 0.58 | M-CSF | 0.67 | G-CSF | 0.70 | FGF-2 | 0.91 |
| IL-3 | 0.57 | FLT-3L | 0.62 | IL-3 | 0.63 | MDC | 0.90 |
| M-CSF | 0.56 | RANTES | 0.62 | IL-15 | 0.63 | IFN-α2 | 0.90 |
| MDC | 0.53 | IL-17F | 0.61 | IL-10 | 0.59 | IL-5 | 0.85 |
| IFN-α2 | 0.51 | IL-18 | 0.61 | IL-8 | 0.59 | IL-17E/IL-25 | 0.84 |
| IL-5 | 0.50 | GROα | 0.61 | TNFα | 0.57 | FLT-3L | 0.77 |
| IL-18 | 0.48 | IL-12p40 | 0.59 | IFNγ | 0.56 | PDGF-AB/BB | 0.73 |
| IL-1RA | 0.45 | IL-10 | 0.55 | FLT-3L | 0.43 | IL-1RA | 0.71 |
| RANTES | 0.45 | PDGF-AA | 0.54 | IL-6 | 0.42 | TGFα | 0.70 |
| IL-10 | 0.45 | IL-8 | 0.53 | IL-17F | 0.38 | IL-4 | 0.66 |
| sCD40L | 0.44 | IFNγ | 0.50 | CRP | 0.36 | PDGF-AA | 0.60 |
| TNFα | 0.42 | IL-17A | 0.46 | GROα | 0.31 | Ferritin | 0.60 |
| TGFα | 0.41 | sCD40L | 0.46 | IL-18 | 0.30 | MCP-1 | 0.54 |
| PDGF-AA | 0.40 | MDC | 0.46 | IL-12p40 | 0.28 | Fractalkine | 0.53 |
| IL-8 | 0.36 | MIP-1α | 0.44 | sCD40L | 0.24 | TNFα | 0.50 |
| MIP-1α | 0.35 | IL-1RA | 0.43 | MIP-1β | 0.20 | VEGF-A | 0.50 |
| PDGF-AB/BB | 0.35 | PDGF-AB/BB | 0.42 | RANTES | 0.15 | RANTES | 0.49 |
| IL-17E/IL-25 | 0.35 | IFN-α2 | 0.40 | MCP-3 | 0.07 | IL-8 | 0.49 |
| IFNγ | 0.34 | IL-12p70 | 0.38 | PDGF-AA | 0.07 | IL-27 | 0.43 |
| GROα | 0.30 | FGF-2 | 0.35 | Fractalkine | -0.04 | M-CSF | 0.39 |
| IL-12p70 | 0.29 | TNFα | 0.35 | IL-22 | -0.07 | IL-9 | 0.39 |
| FGF-2 | 0.29 | MCP-1 | 0.34 | Eotaxin | -0.10 | IL-1β | 0.28 |
| MCP-1 | 0.27 | IL-6 | 0.34 | IL-17A | -0.12 | IL-10 | 0.28 |
| IL-2 | 0.26 | TGFα | 0.33 | IL-13 | -0.15 | GM-CSF | 0.27 |
| IL-27 | 0.24 | IL-4 | 0.33 | IL-12p70 | -0.17 | MCP-3 | 0.19 |
| IL-4 | 0.23 | Eotaxin | 0.33 | TNFβ | -0.18 | IL-3 | 0.16 |
| Eotaxin | 0.23 | IL-5 | 0.32 | IL-2 | -0.21 | MIP-1β | 0.16 |
| IL-7 | 0.21 | IL-2 | 0.31 | IL-4 | -0.21 | Eotaxin | 0.16 |
| GM-CSF | 0.21 | G-CSF | 0.31 | IL-1α | -0.26 | IL-15 | 0.13 |
| EGF | 0.21 | IL-1α | 0.28 | IFN-α2 | -0.33 | IL-1α | 0.05 |
| G-CSF | 0.19 | GM-CSF | 0.28 | IL-7 | -0.34 | IL-7 | 0.01 |
| IL-6 | 0.19 | IL-7 | 0.25 | PDGF-AB/BB | -0.39 | MIP-1α | 0.00 |
| CRP | 0.17 | IL-1β | 0.24 | MDC | -0.40 | IL-13 | -0.02 |
| VEGF-A | 0.17 | EGF | 0.22 | Ferritin | -0.43 | IFNγ | -0.08 |
| IL-17A | 0.15 | VEGF-A | 0.21 | MCP-1 | -0.43 | IL-12p70 | -0.09 |
| IL-9 | 0.14 | IL-9 | 0.18 | EGF | -0.44 | IL-18 | -0.10 |
| IL-1β | 0.12 | CRP | 0.17 | VEGF-A | -0.45 | TNFβ | -0.12 |
| MIP-1β | 0.11 | MIP-1β | 0.17 | IL-5 | -0.46 | IL-2 | -0.12 |
| IL-17F | 0.09 | IL-27 | 0.14 | TGFα | -0.53 | IL-6 | -0.17 |
| IL-22 | 0.06 | IL-22 | 0.14 | IL-27 | -0.53 | **MIG/CXCL9** | **-0.22** |
| MCP-3 | 0.04 | IL-13 | 0.12 | GM-CSF | -0.59 | G-CSF | -0.25 |
| IL-13 | 0.02 | IL-17E/IL-25 | 0.11 | MIP-1α | -0.61 | IL-22 | -0.31 |
| TNFβ | -0.01 | TNFβ | 0.07 | IL-17E/IL-25 | -0.63 | GROα | -0.39 |
| IL-1α | -0.01 | MCP-3 | -0.15 | IL-9 | -0.74 | IL-17F | -0.45 |
| Ferritin | -0.04 | Fractalkine | -0.23 | IL-1β | -0.81 | CRP | -0.53 |
| Fractalkine | -0.21 | Ferritin | -0.29 | FGF-2 | -0.85 | IL-17A | -0.53 |

Pearson r correlation coefficients were calculated between changes in levels of IP-10 with changes in levels of other cytokines between day 3 and baseline. Correlations in each subset of patients are displayed in descending order.

*Recovered to an absolute lymphocyte count of at least 1.0

### Table 3: Secondary Infections following infliximab-abda therapy

| Patients | Diagnosis | Diagnostic certainty | Organism(s) if isolated | Time to diagnosis (days) | Time to treatment (days) | Duration of antimicrobial therapy (days) | Death due to secondary infection(s) |
|---|---|---|---|---|---|---|---|
| **1** | VAP | Suspected | NA | 5 | 5 | 6 | No |
| **2** | VAP | Confirmed | Pseudomonas aeruginosa | 3 | 4 | 5 | NA |
| **3** | CAPA | Probable | Aspergillus fumigatus | 3 | 3 | 41 | NA |
| | CMV viremia | Confirmed | CMV | 25 | NA | NA | |
| | VAP | Confirmed | Acinetobacter baumannii | 26 | 32 | 9 | |
| **4** | VAP | Confirmed | MRSA, MDR-Klebsiella | 2 | 3 | 14 | Confirmed |
| | HSV pneumonitis | Confirmed | HSV | At autopsy | NA | NA | |
| **5** | VAP | Suspected | NA | 12 | 13 | 15 | NA |
| | Tracheostomy site SSTI | Confirmed | Polymicrobial | 21 | 25 | 6 | |
| | CMV viremia | Confirmed | CMV | 24 | NA | NA | |
| **6** | VAP | Confirmed | Polymicrobial | 1 | 1 | 9 | Suspected |
| | CAPA | Probable | NA | 6 | 6 | 3 | |
| **7** | VAP | Suspected | NA | 5 | 5 | 22 | NA |
| | Invasive candidiasis | Suspected | NA | 15 | 23 | 7 | |
| **8** | CAP | Confirmed | Streptococcus pneumoniae | 1 | 1 | 6 | Suspected |
| | HSV pneumonitis | Confirmed | HSV-1 | 14 | 16 | 13 | |
| | CAPA | Possible | Aspergillus niger | 35 | 22 | 13 | |

VAP: Ventilator-associated pneumonia; CAPA: COVID-19-associated pulmonary aspergillosis; CMV: Cytomegalovirus; MRSA: Methicillin-resistant Staphylococcus aureus; HSV: Herpes simplex virus; CAP: Community-acquired pneumonia; MDR: Multidrug resistant; SSTI: Skin and soft tissue infection; NA: Not applicable.
